# Placental Klotho responses to supervised prenatal exercise: findings from the PLACENTRAINING study

**DOI:** 10.64898/2026.10.31.26366971

**Authors:** L. Baena-García, P Acosta-Manzano, J.L. López-Cánovas, M de la Flor Alemany, N.M. Viruez-Arenas, E Martínez-Rosales, F. Amaro-Gahete, V.A. Aparicio

**Author notes:** Corresponding author: Elena Martínez-Rosales. Department of Physiology, Faculty of Pharmacy, University of Granada, Granada, Spain. **Clinical trial registration**: ClinicalTrials.gov (NCT02582567; registered 20 October 2015).

## Abstract

**Background:** Regular exercise during pregnancy improves maternal and neonatal outcomes; however, the molecular pathways underlying placental adaptations to prenatal exercise remain poorly understood. Klotho is a cytoprotective protein involved in the regulation of oxidative stress, metabolic homeostasis, and cellular resilience, yet whether placental Klotho concentrations are associated with prenatal exercise has not been previously investigated.

**Aims:** The primary aim of this study was to examine whether participation in a supervised concurrent exercise program during pregnancy is associated with placental Klotho concentrations at birth. Secondary aims were to explore potential fetal sex–specific patterns and associations between placental Klotho and selected biomarkers related to angiogenic and inflammatory pathways.

**Methods:** Pregnant women were allocated to either a supervised concurrent exercise program or usual care. The intervention comprised combined aerobic and resistance training performed three times per week from early–mid pregnancy until birth. Placental tissue was collected at birth, and Klotho concentrations were quantified by enzyme-linked immunosorbent assay (ELISA). Secondary analyses explored potential sex-specific patterns and examined associations between placental Klotho, placental weight, and a panel of placental biomarkers.

**Results:** Seventy-six women were included (exercise: n = 41; control: n = 35). Placental Klotho concentrations were significantly higher in the exercise group, with an adjusted mean difference of approximately 101 pg/mL *(p* = 0.001; partial η² = 0.164). This effect was consistent across fetal sex, with no significant exercise-by-sex interaction. Several placental biomarkers were independently associated with Klotho, suggesting a potential relationship with angiogenic and inflammatory signaling pathways.

**Conclusions:** Participation in a supervised concurrent exercise program during pregnancy is associated with higher placental Klotho concentrations at birth, irrespective of fetal sex. These findings provide novel insight into molecular pathways potentially involved in placental adaptations associated with prenatal exercise and reinforce the concept of the placenta as an exercise-responsive organ during pregnancy.

## 1. Introduction

Regular physical exercise during pregnancy is widely recognized as a key component of maternal–fetal health(1–5). For instance, previous studies have reported enhanced uteroplacental blood flow(6) and more favorable redox balance in physically active pregnant women(7), suggesting that exercise may promote a more efficient placental environment. Building on this, our group has reported that a supervised concurrent exercise program during pregnancy is associated with greater placental and neonatal weight and shorter labor duration, collectively supporting beneficial effects on placental development and function(8).

The placenta plays a pivotal role in regulating nutrient transport, endocrine signaling and redox balance(9), and in mediating maternal and fetal physiological adaptations to exercise(10). Collectively, these findings support the concept of the placenta as an exercise-responsive organ. Nonetheless, the molecular pathways potentially involved in placental adaptations associated with maternal exercise remain poorly understood, with only few studies investigating placental proteins involved in metabolic regulation, oxidative stress, or cellular ageing(10, 11). Identifying molecular pathways associated with these adaptations may improve understanding of how prenatal exercise contributes to maternal–fetal health.

In the search for molecular mediators potentially involved in these adaptations, α-Klotho (hereafter referred to as Klotho) has emerged as a molecule of particular interest. Klotho is a transmembrane and soluble protein with broad endocrine, paracrine and autocrine actions, originally identified as an anti-aging factor(12). It regulates phosphate– vitamin D metabolism and modulates signaling pathways related to oxidative stress and inflammation(13), and has recognized vasculoprotective and renoprotective effects(14, 15). Regular exercise training increases circulating Klotho concentrations in both healthy and clinical populations(16), supporting its role as a potential exerkine mediating systemic benefits of physical exercise. More recently, Klotho expression has also been demonstrated in human placental tissue, and alterations in its placental or circulating levels have been associated with adverse pregnancy outcomes. Reduced placental or circulating Klotho concentrations have been described in obstetric disorders associated with placental insufficiency such as intrauterine growth restriction(17) and preeclampsia(18), suggesting that Klotho could contribute to placental homeostasis. Despite this emerging evidence, the regulation of placental Klotho expression remains poorly understood. In particular, whether placental Klotho concentrations are associated with prenatal exercise has not been previously investigated, representing an important gap in understanding the molecular pathways through which maternal physical activity may influence placental function. We hypothesised that participation in a supervised prenatal exercise program would be associated with higher placental Klotho concentrations.

Therefore, the primary aim of this study was to examine the association between participation in a supervised concurrent exercise program during pregnancy and placental Klotho concentrations. A secondary aim was to assess whether this response differed by fetal sex, given the recognized sexual dimorphism in placental adaptations to maternal stimuli(19).

## 2. Material and methods

### 2.1 Study design and participants

This study corresponds to the PLACENTRAINING project, conducted within the framework of the GESTAFIT quasi-randomized trial (ClinicalTrials.gov: NCT02582567). The full methodology of the GESTAFIT quasi-randomized trial has been previously reported(20). Briefly, 384 pregnant women were initially approached at their first routine obstetric visit (around the 12^th^ gestational week) at the “San Cecilio” and “Virgen de las Nieves” University Hospitals in Granada (southern Spain). Of these, 159 women met the inclusion criteria (Supplementary Table S1) and provided written informed consent. For the specific objectives of the present study, 76 women with available placental samples and clinical data were analyzed (n=35 control, n=41 exercise). A flowchart detailing participant enrollment, allocation, follow-up, and the derivation of the final analytical sample is provided in Supplementary Figure S1. The trial was approved by the Clinical Research Ethics Committee of Granada (Regional Government of Andalusia, Spain; code GESFIT-0448-N-15; approval date: May 19^th^, 2015).

#### 2.1.1. Sample size

The sample size of the parent GESTAFIT trial was calculated a priori to detect differences in gestational weight gain (n = 52)(20). No specific sample size calculation was performed for the placental outcomes analyzed in the present study, as these analyses were exploratory in nature and based on the availability of placental tissue samples.

### 2.2 Randomization and blinding

The GESTAFIT trial was originally conceived as a randomized controlled study(20). However, during recruitment, participants’ preferences regarding participation in supervised exercise precluded strict random allocation. Consequently, a quasi-randomized, preference-informed allocation approach was adopted, as described previously(20). Women who agreed to participate in supervised exercise were assigned to the exercise group, whereas those who declined structured exercise received usual care. To ensure adequate group balance, additional participants were recruited for the control group.

All participants were followed prospectively and completed the same assessment protocol irrespective of group assignment. Staff involved in maternal assessments and laboratory analyses, including placental tissue processing and Klotho quantification, were blinded to group allocation, and all statistical analyses were conducted using anonymised datasets. The overall assessment procedures are summarized in Figure S2.

### 2.3 Exercise intervention

Women in the exercise group completed a supervised, structured concurrent training program (aerobic and resistance exercise) from the 17^th^ gestational week until birth. Sessions were held three times per week, lasted approximately one hour, and were delivered in small groups by exercise professionals with specific expertise in prenatal exercise prescription.

Each session followed a consistent structure, including a gradual warm-up, a central phase consisting of continuous moderate-intensity aerobic exercises, monitored through heart rate and perceived exertion followed by a resistance-training circuit targeting major muscle groups, and a cool-down incorporating mobility and breathing exercises. Intensity targets were adjusted individually to remain within recommended ranges for pregnancy. Attendance was recorded at every session to monitor adherence. Participants who attended at least 75% of the scheduled sessions were classified as compliant and included in per-protocol analyses. Additional details are provided elsewhere (21). During the intervention, women allocated to the exercise group also received seven brief educational talks on healthy pregnancy, including general recommendations on lifestyle and maternal well-being (Appendix A, Supplementary Material).

### 2.4 Measurements

#### 2.4.1 Sociodemographic and lifestyle data

Participants provided baseline information through self-administered questionnaires. The variables collected included maternal age, educational level, working status, smoking habit and cohabitation with a partner. All data were collected prior to the start of the intervention using identical procedures in both study groups.

#### 2.4.2. Previous obstetric history

Information on previous obstetric history was obtained from the official “Pregnancy Health Document”, which is a standardized medical record provided to all pregnant women by the regional health system in Andalusia. This document contains comprehensive obstetric and gynaecological information and was reviewed for research purposes. Data extracted included gravidity, parity, outcomes of previous pregnancies, past births, and relevant gynaecological antecedents. Gestational age was estimated based on the date of the last menstrual period (assuming a 28-day cycle) and corrected by ultrasound measurements when required.

#### 2.4.3. Body composition

Maternal body composition was assessed at the 16^th^ gestational week following standardized procedures. Weight was measured with a scale (InBody R20, Biospace, Seoul, Korea), with participants wearing light clothing and no shoes. Height was recorded using a stadiometer (Seca22, Hamburg, Germany). Body mass index (BMI) at the 16^th^ gestational week was calculated as measured weight (kg) divided by height squared (m²). All measurements were performed by trained personnel using identical protocols in both study groups.

#### 2.4.4. Birth outcomes

Clinical birth-related information was obtained from the partogram and obstetric records, including gestational age at birth, mode of birth (spontaneous vaginal, instrumental, or caesarean section), newborn sex, birthweight, and duration of labor.

### 2.5 Placental sample processing

For each placenta, samples were thawed under controlled conditions, and approximately 100 mg of tissue were obtained from a standardized biopsy location. The tissue was mechanically disrupted on ice using a T-10 basic ULTRA-TURRAX® homogenizer (IKA, Staufen, Germany) in 1 mL of RIPA buffer (ref. R0278; Sigma-Aldrich, Burlington, MA, USA) supplemented with phenylmethylsulfonyl fluoride (PMSF; 1:200, ref. P7626, Sigma) and a protease inhibitor cocktail (PIC; 1:100, ref. P8340, Sigma).

After homogenization, lysates were maintained at 4 °C with gentle rotation and subsequently clarified by centrifugation at 13,000 g for 15 minutes at 4 °C. The supernatant was collected and a small aliquot (10 µL mixed with 10 µL Milli-Q water) was reserved for protein quantification, while the remaining volume was stored at −80 °C until further analysis.

Total protein concentration was estimated using the Bradford method with the Bio-Rad Protein Assay Dye Reagent Concentrate (ref. 5000006; Bio-Rad Laboratories, Hercules, CA, USA), following the manufacturer’s protocol. Standard curves were prepared in duplicate, and each sample was analyzed in triplicate. Absorbance readings were obtained at 595 nm using a Nanoquant Infinite M200 Pro plate reader (Tecan, Zurich, Switzerland). Protein concentrations were calculated using the Beer–Lambert equation and corrected for the corresponding dilution factors. Finally, extracts were diluted to obtain a final working concentration of 2 µg/mL

### 2.6 Klotho determination in placental samples

Placental concentrations were determined using a commercially available enzyme-linked immunosorbent assay (Antibody-Sandwich ELISA) kit (EH4278, Fine Test, Wuhan Fine Biotech Co., Ltd., Wuhan, China), according to the manufactureŕs instructions. The assay has a reported sensitivity of 56.25 pg/ml, and no significant cross-reactivity or interference with Klotho-related proteins has been described by the manufacturer. O.D. absorbance at 450nm was measured in a VANTAstar multi-mode microplate reader (BMG LABTECH, Inc.; Cary, NC, USA), and the intra- and inter-assay coefficient of variation (% CV) were 2.95% and 5.79%, respectively. The samples were analyzed in duplicates, following standardized procedures to ensure analytical consistency.

### 2.7. Placental inflammatory and angiogenic biomarkers

Placental inflammatory and angiogenic biomarkers were measured using procedures previously described(21). Briefly, frozen placental biopsies were homogenized in radioimmunoprecipitation assay (RIPA) buffer supplemented with protease inhibitors. Homogenates were centrifuged, and supernatants were collected and stored at −80 °C until analysis. Total protein concentration was determined using the Bradford assay, and samples were normalized prior to biomarker quantification.

Placental concentrations of fibroblast growth factor 2 (FGF-2), granulocyte colony-stimulating factor (G-CSF), platelet-derived growth factor AA (PDGF-AA), tumor necrosis factor alpha (TNF-α), and monocyte chemoattractant protein-1 (MCP-1) were quantified using a customized multiplex magnetic bead–based immunoassay (Luminex xMAP technology; MilliporeSigma, Darmstadt, Germany), according to the manufacturer’s instructions. Data acquisition was performed on a Luminex 200 analyzer using xPONENT software. Biomarker concentrations were calculated from five-parameter logistic standard curves and expressed relative to total protein content.

### 2.8 Statistical analysis

Descriptive statistics were computed for all study variables. Continuous variables are presented as mean ± standard deviation, and categorical variables as absolute and relative frequencies. Normality was assessed using graphical methods, including histograms and Q–Q plots, together with the inspection of standardized residuals. Placental Klotho concentrations showed the presence of extreme influential values; therefore, a winsorization procedure at ±2.58 standard deviations was applied to reduce the influence of outliers while retaining all observations. This approach improved distributional properties and was considered appropriate for biomarker data.

Between-group differences in participant characteristics were assessed using independent-samples t tests or chi-square tests, as appropriate. To evaluate associations between participation in the exercise program and placental Klotho concentrations, unadjusted group differences were initially examined using a one-way analysis of variance (ANOVA). Subsequently, adjusted analyses were performed using analysis of covariance (ANCOVA), including a priori covariates selected based on biological plausibility and previous literature: maternal age, body mass index measured at week 16 of gestation as an indicator of early-pregnancy adiposity, and mode of birth (cesarean vs vaginal). Estimated marginal means, unstandardized coefficients (B), 95% confidence intervals, p-values, and partial eta squared (η²ₚ) were reported. Sex-stratified analyses were then performed by repeating the adjusted ANCOVA models within strata of fetal sex. Graphical representations (violin plots) were generated to illustrate sex-dependent patterns. Given the relevance of placental weight as a surrogate marker of placental growth and functional capacity, an exploratory analysis was conducted to examine its association with placental Klotho concentrations. Unadjusted associations were first evaluated, followed by adjusted linear models including the same covariates as the primary analysis. These results are presented in **Supplementary Table S2**.

Given the emerging role of Klotho in pathways related to oxidative stress, inflammation, and tissue homeostasis, associations between placental Klotho concentrations and a selected panel of placental inflammatory and angiogenic biomarkers were explored. Each biomarker was initially entered into an unadjusted linear regression model with Klotho as the dependent variable. Biomarkers showing evidence of association were subsequently examined in adjusted models including exercise group as a covariate. These exploratory analyses are detailed in **Supplementary Table S3**. Analyses were conducted including all participants with available placental samples, regardless of adherence to the exercise program. Per-protocol analyses including women attending ≥75% of sessions were performed as secondary analyses. Additional sensitivity analyses further adjusting for smoking status, educational level, and gestational age at birth, as well as analyses using non-winsorized Klotho values, were conducted and are presented in **Supplementary Table S4**.

All analyses were performed using IBM SPSS Statistics (version 20.0), and statistical significance was set at *p* < 0.05.

## 3. Results

A total of 76 pregnant women were included in the analyses (mean age 33.2 ± 4.6 years, BMI 24.8 ± 4.1 kg/m² at 16^th^ gestational week). Sociodemographic and clinical characteristics of the participants are shown in **Table 1**. Most participants lived with a partner, and approximately two-thirds had completed university education. Smoking during early pregnancy was uncommon, and around 60% of women were nulliparous. Regarding birth outcomes, most births were vaginal, with a mean gestational age at birth of 40 weeks. Mean placental weight was approximately 567 g, with slightly higher values observed in the exercise group compared with the control group, although this difference did not reach statistical significance. Adherence to the supervised program was high, with an average attendance of 85%.

**Table 1.** Sociodemographic and clinical characteristic of the study sample (n=76)

| **Maternal outcomes** | **All women (n=76)** | **Control group (n=35)** | **Exercise Group (n=41)** | ***p-value*** |
|---|---|---|---|---|
| **Age, years** | 33.2 (4.6) | 32.89 (4.1) | 33.39 (4.24) | 0.6 |
| **Percentage of attendance (%)** |  |  | 85.2% |  |
| ***Body mass index at the 16<sup>th</sup> gestational week , kg/m<sup>2</sup>*** | 24.8 (4.1) | 24.3 (3.3) | 24.4 (4.1) | 0.8 |
| **Smoking status, n %** | 5 (6.6%) | 4 (11.4) | 1 (2.4) | 0.1 |
| **Living with partner, n %** | 76 (100%) | 35 (100%) | 41 (100%) |  |
| **Educational status, n %** |  |  |  | 0.9 |
| ***Primary or high school*** | 15 (19.7%) | 7 (20) | 8 (19.5) |  |
| ***Specialized training*** | 12 (15.8%) | 4 (11.4) | 8 (19.5) |  |
| ***University degree*** | 49 (64.5%) | 24 (68.6) | 25 (61) |  |
| **Parity, n %** |  |  |  |  |
| ***Nulliparous*** | 47 (61.8%) | 21 (60%) | 26 (63.4%) | 0.7 |
| **Birth outcomes** |  |  |  |  |
| **Type of birth, n %** |  |  |  |  |
| ***Vaginal birth*** | 57 (79.2%) | 26 (74.3) | 31 (75.6) | 0.9 |
| ***Gestational age at birth, weeks*** | 39.6 (1.2) | 39.6 (1.4) | 39.7 (1.1) | 0.5 |
| ***Neonatal outcomes*** |  |  |  |  |
| ***Sex (female), n %*** | 36 (49.3%) | 16 (44.4%) | 20 (55.6%) | 0.9 |
| ***Placental weight, grams*** | 566.9 (91.7) | 544.1 (73.7) | 584.7 (102.3) | 0.06 |
| **Placental proteins** |  |  |  |  |
| ***Klotho, pg/mL*** | 321.1 (125.3) | 267.6 (99.7) | 365.9 (129.6) | **<0.01** |

BMI, body mass index.

### 3.1. Primary analysis: association between exercise-group allocation and placental Klotho concentrations

Placental Klotho concentrations were significantly higher in the exercise group compared with the control. As shown in **Figure 1**, the distribution of Klotho levels was shifted toward higher values in women assigned to the exercise program. After adjusting for maternal age, BMI at 16^th^ gestational week, and mode of birth, placental Klotho concentrations were approximately 101 pg/mL higher in the exercise compared with the control group (B = 100.9, 95% CI 156.9 to 44.9, *p* = 0.001), corresponding to a moderate effect size (partial η² = 0.164). None of the included covariates were independently associated with placental Klotho concentrations.

**Figure 1.**
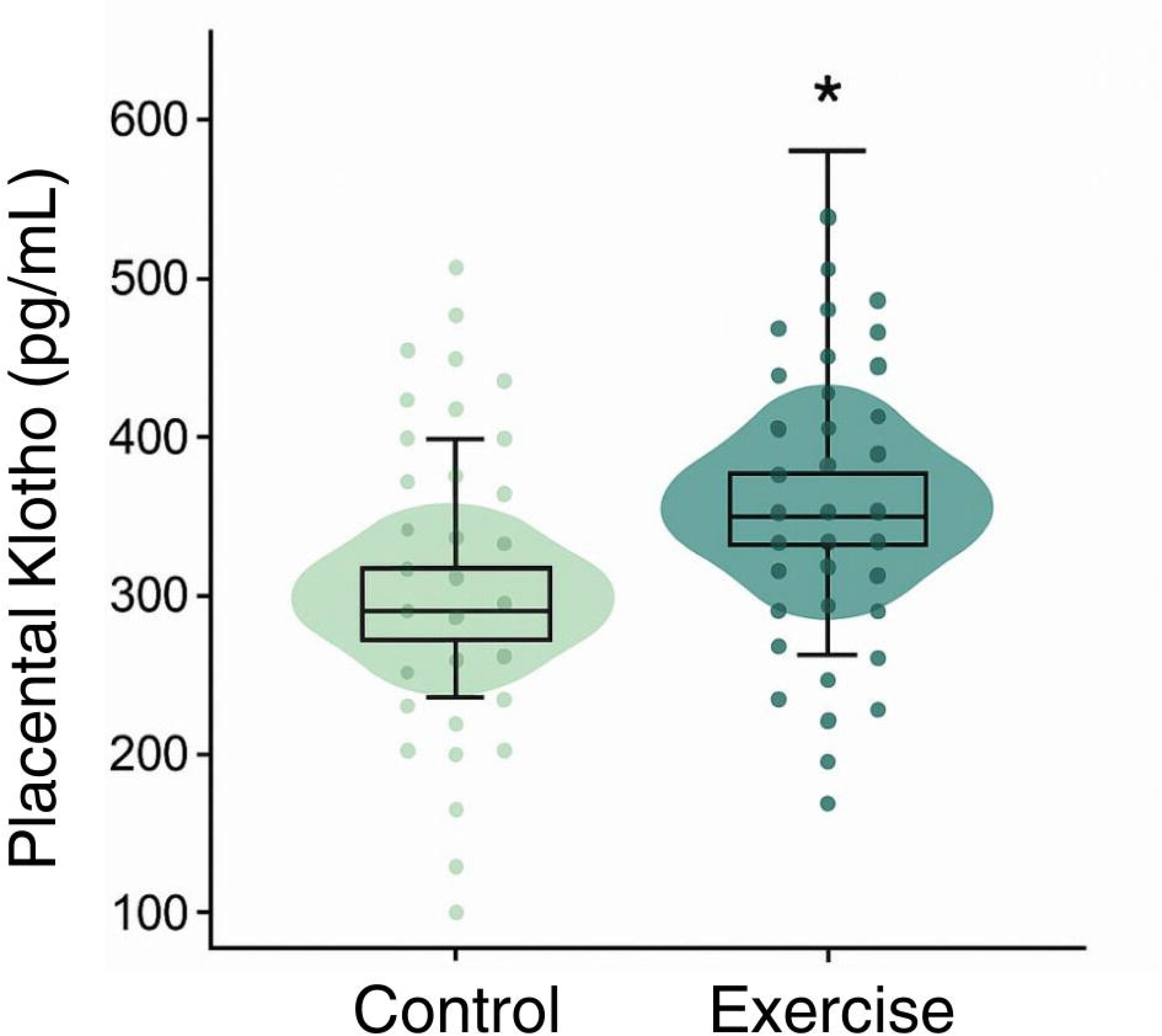
Distribution of placental Klotho concentrations (pg/mL) in the control and exercise groups. Data are presented as mean ± standard deviation. The asterisks (\**p* <0 .05) indicate statistically significant differences.

### 3.2. Sex-stratified analyses

Sex-stratified models revealed a consistent pattern, with higher placental Klotho concentrations in the exercise group for both male and female neonates (**Table 2**). Among male fetuses, adjusted placental Klotho concentrations were higher in the exercise group compared to controls (368.3 ± 27.3 vs. 281.9 ± 28.9 pg/mL), with an adjusted mean difference of 86.4 pg/mL (95% CI 4.7 to 168.2, *p* = 0.039). Similarly, among female fetuses, placental Klotho concentrations were higher in the exercise group (367.2 ± 28.4 vs. 246.7 ± 32.0 pg/mL in controls), corresponding to an adjusted mean difference of 120.5 pg/mL (95% CI 32.3 to 208.7, *p* = 0.009). The direction and magnitude of the association were broadly comparable across sexes, with no evidence that fetal sex materially modified the association between exercise-group allocation and placental Klotho concentrations.

**Table 2.** Placental Klotho levels (pg/mL) by fetal sex and exercise group.

| **Fetal sex** | **Group** | **Adjusted mean Klotho (pg/mL, SE)** | **Mean difference vs. control (95% CI)** | **p-value** |
|---|---|---|---|---|
| **Male fetuses** | Control (n = 17) | 281.86 (28.91) | Reference | — |
|  | Exercise (n = 19) | 368.26 (27.33) | +86.40 (4.65, 168.15) | **0.039** |
| **Female fetuses** | Control (n = 15) | 246.74 (32.03) | Reference | — |
|  | Exercise (n = 19) | 367.19 (28.41) | +120.45 (32.26, 208.65) | **0.009** |

Model adjusted for maternal age, body mass index at the 16<sup>th</sup> gestational week and mode of delivery.

### 3.3. Exploratory analysis: potential maternal and placental determinants of Klotho expression

In exploratory analyses, we first examined the associations between placental Klotho concentrations and a panel of placental inflammatory and angiogenic biomarkers (**Supplementary Table S3**). To reduce the risk of multiple testing and exploratory bias, analyses were restricted to biomarkers with biologically plausible relevance and adequate data quality, defined by sufficient detectability and sample size. Unadjusted linear regression models revealed several biomarkers showing significant associations with Klotho, including FGF-2, G-CSF, PDGF-AA, TNF-α and MCP-1. When each biomarker was subsequently examined in adjusted models controlling for exercise group, FGF-2, G-CSF, PDGF-AA, and TNF-α remained independently associated with placental Klotho concentrations, whereas the association with MCP-1 was no longer statistically significant.

We then explored the potential association between placental weight and placental Klotho concentrations (**Table S2**). Placental weight showed a small and non-significant inverse association with Klotho concentrations in unadjusted analysis. However, in adjusted models including maternal age, BMI, and mode of birth, placental weight emerged as an independent predictor, such that each additional gram of placental tissue was associated with slightly lower Klotho concentration (B = –0.52, 95% CI –0.85 to –0.19, p = 0.002). Despite statistical significance, the magnitude of this association was modest.

Additional potential covariates were examined in sensitivity analyses, including gestational age at birth, and smoking status. These variables were not retained in the primary models, as their inclusion did not materially alter the magnitude or statistical significance of the association between exercise-group allocation and placental Klotho concentrations. Additional potential covariates were examined in sensitivity analyses, including gestational age at birth, smoking status, and educational level (**Supplementary Table S4**). These variables were not retained in the primary models, as their inclusion did not materially alter the magnitude or statistical significance of the association between exercise-group allocation and placental Klotho concentrations.

## 4. Discussion

In this study, women who participated in a supervised concurrent exercise program during pregnancy exhibited higher concentrations of placental Klotho at term birth. The magnitude of this difference suggests a potentially meaningful association between prenatal exercise and Klotho-related pathways involved in the regulation of oxidative stress, angiogenic signaling, and inflammatory responses. These findings broaden the current understanding of how structured exercise may influence placental biology, indicating that exercise-related adaptations extend beyond vascular and metabolic domains to include proteins involved in cytoprotection and cellular resilience.

To the best of our knowledge, no previous studies have examined whether maternal exercise modulates Klotho expression in the human placenta. While prior research has documented multiple placental adaptations to exercise, including increased capillary volume and exchange surface(22), enhanced mitochondrial function(23), and alterations in angiogenic balance or villous structure in physically active women(22, 24, 25), evidence regarding the regulation of proteins directly involved in redox balance and cellular longevity has remained limited. In this context, our findings extend existing evidence and align with human studies showing that regular exercise increases Klotho levels in circulating compartments and, in some cases, in peripheral tissues such as skeletal muscle(26–28). Collectively, these data support the biological plausibility that exercise during gestation may activate molecular pathways with cytoprotective potential in placental cells.

The sex-stratified analyses further refine this interpretation. Although women assigned to the exercise intervention displayed higher placental Klotho concentrations regardless of fetal sex, the magnitude of the association appeared numerically greater in female placentas. At the same time, baseline Klotho concentrations tended to be higher in males, a pattern that is broadly consistent with the recognized sexual dimorphism in placental physiology(29). Specifically, male placentas often exhibit higher metabolic activity and heightened stress responsiveness, whereas female placentas are thought to display greater adaptive plasticity in response to favorable or adverse maternal conditions(30). Our results suggest that placental Klotho may follow a sex-dimorphic baseline pattern, while its responsiveness to maternal exercise is largely conserved across sexes. Importantly, the absence of a significant sex-by-group interaction, together with overlapping confidence intervals, indicates that the association between exercise-group allocation and placental Klotho concentrations appears broadly sex-independent. Nevertheless, the modest differences observed between male and female strata may point to subtle sex-specific regulatory features that warrant confirmation in larger, adequately powered studies.

Exploratory analyses provide additional insight into factors potentially linked to placental Klotho regulation. Placental weight emerged as a modest but independent predictor of placental Klotho concentrations. As placental weight is commonly used as a surrogate marker of placental growth and function, this modest inverse association may reflect subtle differences in placental maturation or efficiency and should be interpreted cautiously as hypothesis-generating.

More consistent associations were observed between placental Klotho concentrations and placental biomarkers (full regression models are provided in Supplementary Table S3). The positive correlations with FGF-2 and PDGF-AA are consistent with Klotho’s proposed role in vascular and tissue homeostasis(12), whereas the association with TNF-α may reflect shared regulatory pathways or compensatory responses to inflammatory signaling, given Klotho’s involvement in the modulation of inflammatory and oxidative stress–related pathways(12, 31). In the same study population and exercise protocol(32), our group has previously reported that prenatal exercise modulates key placental inflammatory and angiogenic markers, including higher G-CSF levels and lower epidermal growth factor and interleukin-1 receptor antagonist concentrations in the exercise group, as well as sex-specific effects on granulocyte colony-stimulating factor, tumor necrosis factor alpha, and interleukin-6(21).

Within this context, the observed associations between placental Klotho and inflammatory and angiogenic markers suggest that Klotho does not act in isolation but is integrated within a broader placental signaling milieu shaped by maternal exercise. These findings are consistent with our previous report in the same cohort showing exercise-induced modulation of placental inflammatory and angiogenic markers.

Several biological mechanisms may plausibly contribute to the higher placental Klotho concentrations observed in women who exercise during pregnancy. Klotho is a key regulator of oxidative stress, inflammatory signaling and cellular metabolism, and its expression increases under conditions of improved mitochondrial function and reduced cellular stress(12). Prenatal exercise has been proposed to enhance placental antioxidant capacity and to support mitochondrial adaptations, as suggested by recent experimental and review studies(33–35), while also modulating cytokine and growth factor profiles toward a more anti-inflammatory and pro-angiogenic milieu(21).

Furthermore, exercise-induced increases in uteroplacental perfusion may expose placental cells to altered mechanical forces, such as shear stress, which are recognized regulators of endothelial and trophoblast biology(36, 37). Although direct evidence linking shear stress to Klotho regulation in human placental tissue is currently lacking, these converging pathways support a biologically coherent framework in which prenatal exercise may create a placental microenvironment conducive to Klotho upregulation. Given the central role of Klotho in oxidative balance, vascular health and cellular resilience, these results provide a novel perspective on the molecular pathways through which prenatal exercise may contribute to placental and fetal well-being. Future studies should aim to elucidate the upstream mechanisms regulating placental Klotho expression in placental tissue and determine whether exercise-induced modulation of placental Klotho translates into clinically meaningful benefits for both the mother and offspring.

### Limitations and strengths

Some limitations should be acknowledged. The quasi-randomized design, although pragmatic, may be associated with residual selection bias related to participants’ exercise preferences, despite prospective follow-up and generally comparable baseline characteristics between groups. This limitation reflects a common methodological and ethical barrier in antenatal exercise research, where participant preferences and feasibility constraints frequently preclude full randomization(38). The sample size, while sufficient to detect between-group differences in placental Klotho concentrations, limits the precision of sex-specific estimates and reduces statistical power to formally test interaction effects. In addition, the absence of direct assessment of placental perfusion and shear stress precludes definitive conclusions regarding the mechanistic pathways underlying the observed associations. Despite these limitations, the present study has important strengths. It is the first study, to our knowledge, to examine the influence of a prenatal exercise program on placental Klotho concentration, providing novel molecular insight into how maternal physical activity may shape placental biology. The use of a supervised and well-characterized concurrent exercise intervention with objective monitoring of adherence (85%), enhances internal validity. Finally, the incorporation of sex-stratified analyses adds valuable information regarding the potential role of fetal sex in placental molecular adaptations to maternal exercise.

## 5. Conclusion

A supervised concurrent exercise program during pregnancy is associated with higher placental Klotho concentrations, with findings across both male and female fetuses. These findings support the rationale for further translational research aimed at determining whether Klotho could serve as a biomarker of placental responsiveness to lifestyle interventions, or potentially contribute to strategies for optimizing prenatal care.

## Data Availability

The datasets used and/or analyzed during the current study are available from the corresponding author upon reasonable request.

## Authorś contributions

LBG contributed to conceptualization, methodology, data curation, formal analysis, visualization, investigation, and writing (original draft); PAM contributed to methodology, investigation, and placental sample processing; JLLC contributed to methodology and laboratory analyses; MFA and EMR contributed to investigation, placental sample processing, and data curation; NMVA contributed to investigation; FAG contributed to methodology and laboratory analyses; VAA contributed to conceptualization, methodology, supervision, and resources. All authors contributed to writing (review and editing), read and approved the final version of the manuscript, and agree with the order of presentation of the authors.

## Acknowledgements

We thank all women who participated in the GESTAFIT and PLACENTRAINING studies. AI-assisted editing tools were used to enhance clarity and grammar. The authors retain full responsibility for the scientific integrity of the manuscript. The placental analyses included in this study were supported by the PLACENTRAINING project, funded through the FEDER-UGR23 funding call (European Regional Development Fund – University of Granada program; grant number C-EXP-336-UGR23). JLLC and EMR were supported by the Juan de la Cierva Grant (JDC2022-049274-I & JDC2023-051490-I, respectively) founded by MCIN/AEI/10.13039/501100011033 and by “’ESF+”. Participant recruitment, follow-up, and the exercise intervention were conducted within the GESTAFIT trial (ClinicalTrials.gov: NCT02582567). The funders had no role in study design, data collection, analysis, interpretation, or manuscript preparation.

Supplemental Digital Content 1. docx

## References

1. Ruiz JR, Perales M, Pelaez M, Lopez C, Lucia A, Barakat R. Supervised exercise-based intervention to prevent excessive gestational weight gain: A randomized controlled trial. Mayo Clin Proc. 2013;88(12):1388–97.

2. Davenport MH, Ruchat SM, Poitras VJ, et al. Prenatal exercise for the prevention of gestational diabetes mellitus and hypertensive disorders of pregnancy: A systematic review and meta-analysis. Br J Sports Med. 2018;52(21):1367–75.

3. Barakat R, Pelaez M, Cordero Y, et al. Exercise during pregnancy protects against hypertension and macrosomia: Randomized clinical trial. Am J Obstet Gynecol. 2016;214(5):649.e1–649.e8.

4. Giles C, Johnston R, Kubler J, Spathis J, Beetham K. The effects of aerobic and resistance exercise on blood pressure in uncomplicated and at risk pregnancies: A systematic review and meta-analysis. Women’s Heal. 2023;19:17455057231183572.

5. Marín-Jiménez N, Baena-García L, Coll-Risco I, Flor-Alemany M, Castro-Piñero J, Aparicio VA. Influence of a Concurrent Exercise Training Program on Health-Related Quality of Life During Advanced Pregnancy: The GESTAFIT Project. Sports Health. 2024;16(4):518–26.

6. Clapp JF. The effects of maternal exercise on fetal oxygenation and feto-placental growth. European Journal of Obstetrics and Gynecology and Reproductive Biology. 2003. p. 80–5.

7. Ramírez-Vélez R, Aguilar de Plata AC, Escudero MM, et al. Influence of regular aerobic exercise on endothelium-dependent vasodilation and cardiorespiratory fitness in pregnant women. J Obstet Gynaecol Res. 2011;37(11):1601–8.

8. Baena-García L, de la Flor-Alemany M, Coll-Risco I, Reoyo OR, Aranda P, Aparicio VA. A concurrent prenatal exercise program increases neonatal and placental weight and shortens labor: The GESTAFIT project. Scand J Med Sci Sports. 2023;33(4):465–74.

9. Sanchez-Aranguren L, Nadeem S. Bioenergetics adaptations and redox homeostasis in pregnancy and related disorders. Mol Cell Biochem. 2021;476(11):4003–18.

10. Adamo KB, Goudreau AD, Corson AE, MacDonald ML, O’Rourke N, Tzaneva V. Physically active pregnancies: Insights from the placenta. Physiol Rep. 2024;12(11):e16104.

11. Jevtovic F, Wisseman BL, Jahan F, et al. Maternal exercise alters placental proteome in an exercise mode-specific manner. Am J Physiol Endocrinol Metab [Internet]. 2025 [cited 2025 Dec 19];329(6) Available from: https://pubmed.ncbi.nlm.nih.gov/41187954/. doi:10.1152/AJPENDO.00052.2025.

12. Kuro-o M. Klotho and aging. Biochim Biophys Acta - Gen Subj. 2009;1790(10):1049–58. [cited 2025 Dec 3 ] Available from: https://pubmed.ncbi.nlm.nih.gov/19230844/.

13. Quarles LD. Fibroblast growth factor 23 and α-Klotho co-dependent and independent functions. Curr Opin Nephrol Hypertens. 2019;28(1):16–25.

14. Kanbay M, Brinza C, Ozbek L, et al. The association between klotho and kidney and cardiovascular outcomes: a comprehensive systematic review and meta-analysis. Clin Kidney J [Internet]. 2024 [cited 2025 Dec 10];17(9) Available from: https://pubmed.ncbi.nlm.nih.gov/39281418/. doi:10.1093/CKJ/SFAE255.

15. Drew DA, Katz R, Kritchevsky S, et al. Association between Soluble Klotho and Change in Kidney Function: The Health Aging and Body Composition Study. J Am Soc Nephrol. 2017;28(6):1859.

16. Amaro-Gahete FJ, de-la-O A, Jurado-Fasoli L, Ruiz JR, Castillo MJ, Gutiérrez Á. Role of Exercise on S-Klotho Protein Regulation: A Systematic Review. Curr Aging Sci. 2018;11(2):100–7.

17. Zeng G, Lian J, Shen J, Shi Y. Expression and clinical significance of Klotho protein in serum, umbilical cord blood, and placenta of pregnant women with intrauterine growth restriction. Front Pediatr [Internet]. 2025 [cited 2025 Dec 10];13 Available from: https://pubmed.ncbi.nlm.nih.gov/40831795/. doi:10.3389/FPED.2025.1611877.

18. Cecati M, Giannubilo SR, Saccucci F, et al. Potential Role of Placental Klotho in the Pathogenesis of Preeclampsia. Cell Biochem Biophys 2015 741. 2015;74(1):49–57.

19. Cvitic S, Longtine MS, Hackl H, et al. The human placental sexome differs between trophoblast epithelium and villous vessel endothelium. PLoS One [Internet]. 2013 [cited 2025 Dec 19];8(10) Available from: https://pubmed.ncbi.nlm.nih.gov/24205377/. doi:10.1371/JOURNAL.PONE.0079233.

20. Aparicio VA, Ocón O, Padilla-Vinuesa C, et al. Effects of supervised aerobic and strength training in overweight and grade I obese pregnant women on maternal and foetal health markers: the GESTAFIT randomized controlled trial. BMC Pregnancy Childbirth. 2016;16(1):290.

21. Acosta-Manzano P, Flor-Alemany M, Martínez-González LJ, et al. The effects of a supervised exercise training program during pregnancy on placental cytokines, and the potential role of fetal sex and maternal weight status. J Sport Heal Sci. 2025;101082.

22. Jackson MR, Gott P, Lye SJ, Ritchie JW, Clapp JF. The effects of maternal aerobic exercise on human placental development: placental volumetric composition and surface areas. Placenta. 1995;16(2):179–91.

23. Aparicio VA, Baena-García L, Flor-Alemany M, et al. Differences in maternal and neonatal cardiometabolic markers and placenta status by foetal sex. The GESTAFIT project. Women’s Heal [Internet]. 2022;18 doi:10.1177/17455057221117976.

24. Hardy DB, Mu X, Marchiori KS, Mottola MF. Exercise in Pregnancy Increases Placental Angiogenin without Changes in Oxidative or Endoplasmic Reticulum Stress. Med Sci Sports Exerc. 2021;53(9):1846–54.

25. Bergmann A, Zygmunt M, Clapp JF. Running throughout pregnancy: Effect on placental villous vascular volume and cell proliferation. Placenta. 2004;25(8–9):694–8.

26. Corrêa H de L, Raab ATO, Araújo TM, et al. A systematic review and meta-analysis demonstrating Klotho as an emerging exerkine. Sci Rep. 2022;12(1):17587.

27. Arroyo E, Troutman AD, Moorthi RN, Avin KG, Coggan AR, Lim K. Klotho: An Emerging Factor With Ergogenic Potential. Front Rehabil Sci [Internet]. 2022 [cited 2025 Dec 3];2 Available from: https://pubmed.ncbi.nlm.nih.gov/36188832/. doi:10.3389/FRESC.2021.807123.

28. Amaro-Gahete FJ, De-la-O A, Jurado-Fasoli L, et al. Exercise training increases the S-Klotho plasma levels in sedentary middle-aged adults: A randomised controlled trial. The FIT-AGEING study. J Sports Sci. 2019;37(19):2175–83.

29. Clifton VL. Review: Sex and the Human Placenta: Mediating Differential Strategies of Fetal Growth and Survival. Placenta. 2010;31(SUPPL.):S33–9.

30. Clifton VL. Review: Sex and the human placenta: mediating differential strategies of fetal growth and survival. Placenta [Internet]. 2010 [cited 2025 Dec 10];31 Suppl(SUPPL.) Available from: https://pubmed.ncbi.nlm.nih.gov/20004469/. doi:10.1016/J.PLACENTA.2009.11.010.

31. Prud’homme GJ, Wang Q. Anti-Inflammatory Role of the Klotho Protein and Relevance to Aging. Cells 2024, Vol 13,. 2024;13(17):1413.

32. Aparicio VA, Ocón O, Padilla-Vinuesa C, et al. Effects of supervised aerobic and strength training in overweight and grade I obese pregnant women on maternal and foetal health markers: the GESTAFIT randomized controlled trial. BMC Pregnancy Childbirth. 2016;16(1):290.

33. Chae SA, Son JS, Du M. Prenatal exercise in fetal development: a placental perspective. FEBS J. 2022;289(11):3058–71.

34. Alizadeh Pahlavani H. Possible roles of exercise and apelin against pregnancy complications. Front Endocrinol (Lausanne) [Internet]. 2022 [cited 2025 Dec 10];13 Available from: https://pubmed.ncbi.nlm.nih.gov/36093083/. doi:10.3389/FENDO.2022.965167.

35. Pahlavani HA, Laher I, Weiss K, Knechtle B, Zouhal H. Physical exercise for a healthy pregnancy: the role of placentokines and exerkines. J Physiol Sci. 2023;73(1):30.

36. Lecarpentier E, Bhatt M, Bertin GI, et al. Computational Fluid Dynamic Simulations of Maternal Circulation: Wall Shear Stress in the Human Placenta and Its Biological Implications. PLoS One. 2016;11(1):e0147262.

37. Morley LC, Beech DJ, Walker JJ, Simpson NAB. Emerging concepts of shear stress in placental development and function. Mol Hum Reprod. 2019;25(6):329–39.

38. Kehler AK, Heinrich KM. A selective review of prenatal exercise guidelines since the 1950s until present: Written for women, health care professionals, and female athletes. Women and Birth. 2015;e93–8.

