## Supplementary material for "Placental Klotho responses to supervised prenatal exercise: findings from the PLACENTRAINING study": Suplementary Material

**Table of contents**

**Appendix A.** Talks provided to pregnant women

**Table S1**. Inclusion and exclusion criteria in the GESTAFIT project

**Table S2.** Exploratory association between placental weight and placental Klotho concentrations.

**Table S3.** Exploratory analyses examining associations between placental biomarkers and placental Klotho concentrations.

**Table S4.** Sensitivity analyses for the association between participation in the exercise program and placental Klotho concentrations

**Figure S1.** Flowchart: enrolment, allocation, follow-up and analyses.

**Figure S2.** General assessment procedures.

.

**Appendix A. Talks provided to pregnant women**

During the intervention, the research team gave 7 lectures to pregnant women from both groups (exercise and control group) about: 1) the benefits of physical exercise for a better pregnancy, prevention and treatment of cardiovascular diseases and excessive weight gain; 2) ergonomic advice, exercises to perform at home and strategies to increase their daily physical activity levels; 3) the benefits of the Mediterranean Diet and adequate nutritional habits during pregnancy; 4) how to avoid toxics and chemicals during the pregnancy and breastfeeding; 5) pregnancy, postpartum and sex; 6) physical and mental preparation for the labour, what to expect; 7) nutritional education towards breastfeeding. We also used these conferences to maintain control group fidelity until the end of the program.

**Table S1.** Inclusion and exclusion criteria in the GESTAFIT project

***Inclusion criteria***

- Pregnant women aged 25-40 years old with a normal pregnancy course.

- Answering “no” to all questions on the PARmed-X for pregnancy.

- Being able to walk without assistance.

- Being able to read and write properly.

- Informed consent: Being capable and willing to provide written consent.

***Exclusion criteria***

- Having acute or terminal illness.

- Having malnutrition.

- Being unable to conduct tests for assessing physical fitness or exercise during pregnancy.

- Having severe pregnancy risk factors (e.g., hypertension, type 2 diabetes).

- Having a multiple pregnancy.

- Having chromosopathy or foetal malformations.

- Having uterine growth restriction.

- Having foetal death.

- Having upper or lower extremity fracture in the past 3 months.

- Suffering neuromuscular disease or presence of drugs affecting neuromuscular function.

- Being registered in another exercise program.

- Performing more than 300 minutes of at least moderate physical activity per week.

- Being engaged in another physical exercise program

- Being unwilling either to complete the study requirements or to be randomized into the control or exercise group.

**Table S2. Exploratory association between placental weight and placental Klotho concentrations.**

| **Variable** | **ρ (unadjusted)** | **p-value (unadjusted)** | **B (adjusted)** | **95% CI for B** | **p-value (adjusted)** | **Partial η²** |
| --- | --- | --- | --- | --- | --- | --- |
| Placental weight (g) | –0.199 | 0.098 | –0.519 | –0.846 to –0.192 | 0.002 | 0.146 |

**Model adjusted for:** maternal age (week 16), BMI (week 16), mode of birth (cesarean/vaginal), and intervention group (intention-to-treat). ρ = Spearman’s rank correlation coefficient.

**Table S3. Exploratory analyses examining associations between placental biomarkers and placental Klotho concentrations**

|  | **Klotho protein (pg/mL)** | | | | | | | |
| --- | --- | --- | --- | --- | --- | --- | --- | --- |
|  | **Unadjusted** | | | | **Exercise-adjusted** | | | |
| **Placental marker** | **β** | **B** | **SE** | **P** | **β** | **B** | **SE** | **P** |
| TNF-α | 0.260 | 1.967 | 0.850 | **0.023** | 0.321 | 2.430 | 0.782 | **0.003** |
| FGF-2 | 0.471 | 0.032 | 0.007 | **<0.001** | 0.418 | 0.028 | 0.007 | **<0.001** |
| G-CSF | 0.518 | 1.127 | 0.217 | **<0.001** | 0.442 | 0.962 | 0.226 | **<0.001** |
| PDGF-AA | 0.581 | 3.981 | 0.649 | **<0.001** | 0.561 | 3.843 | 0.595 | **<0.001** |
| MCP-1 | 0.157 | 0.118 | 0.086 | 0.175 | 0.188 | 0.141 | 0.080 | 0.082 |

IL-1β, interleukin-1 beta; IL-1ra, interleukin-1 receptor antagonist; IL-6, interleukin-6; IL-8, interleukin-8; IL-10, interleukin-10; IFN-γ, interferon-gamma; TNF-α, tumor necrosis factor-alpha; TNF-β, tumor necrosis factor-beta; BDNF, brain-derived neurotrophic factor; EGF, epidermal growth factor; FGF-2, fibroblast growth factor-2; G-CSF, granulocyte colony-stimulating factor; GM-CSF, granulocyte-macrophage colony-stimulating factor; PDGF-AA, platelet-derived growth factor-AA; MCP-1, monocyte chemoattractant protein-1; VEGF, vascular endothelial growth factor. SE: Standard Error; β, standardized regression coefficient; B, nonstandardized regression coefficient.

**Table S4. Sensitivity analyses for the association between participation in the exercise program and placental Klotho concentrations**

| **Model** | **B (pg/mL)** | **95% CI** | **p-value** | | **Partial η²** | |
| --- | --- | --- | --- | --- | --- | --- |
| Primary adjusted model | 100.9 | 44.9 to 156.9 | | 0.001 | | 0.164 |
| + smoking + educational level | 94.7 | 34.2 to 155.1 | | 0.003 | | 0.139 |
| + gestational age at birth, smoking and educational level | 99.7 | 34.2 to 165.3 | | 0.004 | | 0.147 |

**Models adjusted for**: maternal age, BMI at 16 weeks’ gestation, mode of delivery, smoking status, educational level and gestational age at birth

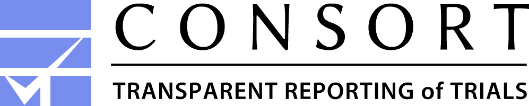

Excluded (n=162)

  Did not answer the phone (n=52)

  Declined to participate (n=110)

Participants assessed for eligibility (n=384)

### Enrolment

Invited for 1^st^ assessment (n=222)

Excluded (n=63)

  Not meeting inclusion criteria (n=5)

	Did not come to 1^st^ assessment (n=58)

Total participants (n=159)

### Allocation

Allocated to exercise intervention (n=72)

Allocated to control (n=87)

### Follow-Up

Lost to follow-up (n=10)

-Did not come at week of gestation 34 (n=33)

-No data was taken at delivery (n=10; women did not contact us or attended a private hospital, or midwives did not collect data/samples)

Lost to follow-up (n=0)

-Did not come at week of gestation 34 (n=2)

-Discontinued intervention (did not reach the 75% of attendance in the exercise program) (n=22)

### Analyses

When appropriate and prior to analyses, censored data was imputed according to previous evidence

**Participants included in placental analyses (birth, n=35)**

Placental Klotho measured (n = 40)

– Excluded due to missing covariate data (n = 5)

When appropriate and prior to analyses, censored data was imputed according to previous evidence

**Participants included in placental analyses (birth, n=41)**

Exercise group (≥75% attendance)

Placental Klotho measured (n = 42)

– Excluded due to missing covariate data (n = 1)

**Figure S1.** Flowchart: enrolment, allocation, follow-up and analyses.

**Second assessment (week 34)**

First day evaluation:

- Sociodemographic and clinical data
- Weight status
- Physical Fitness
- Nutritional assessment

Second day evaluation:

- Accelerometers collection and review of questionnaires
- Placental tissue processing and protein extraction
- Determination of placental Klotho by ELISA
- Quantification of placental inflammatory and angiogenic biomarkers

**Recruitment at Hospital**

- Inclusion and exclusion criteria
- Informed consent
- Self-reported pre-pregnancy weight
- Weight and height measures

**Laboratory analyses**

**Birth**

**12**

**16**

**34**

**Week of gestation**

- Collection of placental tissue samples
- Assessment of birth outcomes (gestational age, mode of birth, neonatal sex, birthweight)
- Measurement of placental weight

**First assessment (week 16)**

First day evaluation:

- Sociodemographic and clinical data
- Weight status
- Physical Fitness
- Nutritional assessment
- Accelerometry

Second day evaluation:

- Accelerometers collection and review of questionnaires

**START OF THE INTERVENTION PROGRAM**

**Figure S2.** General assessment procedures.
